# Reproducibility of radiomic features of the brain on ultrahigh-resolution MRI at 7 Tesla: a comparison of different segmentation techniques

**DOI:** 10.1101/2024.06.24.24308597

**Authors:** Julian H. Klinger, Doris Leithner, Sungmin Woo, Michael Weber, H. Alberto Vargas, Marius E. Mayerhoefer

## Abstract

**Objectives:** To determine the impact of segmentation techniques on radiomic features extracted from ultrahigh-field (UHF) MRI of the brain.

**Materials and Methods:** Twenty-one 7T MRI scans of the brain, including a 3D magnetization-prepared two rapid acquisition gradient echo (MP2RAGE) T1-weighted sequence with an isotropic 0.63 mm³ voxel size, were analyzed. Radiomic features (histogram, texture, and shape; total n=101) from six brain regions -cerebral gray and white matter, basal ganglia, ventricles, cerebellum, and brainstem-were extracted from segmentation masks constructed with four different techniques: the iGT (reference standard), based on a custom pipeline that combined automatic segmentation tools and expert reader correction; the deep-learning algorithm Cerebrum-7T; the Freesurfer-v7 software suite; and the Nighres algorithm. Principal components (PCs) were calculated for histogram and texture features. To test the reproducibility of radiomic features, intraclass correlation coefficients (ICC) were used to compare Cerebrum-7, Freesurfer-v7, and Nighres to the iGT, respectively.

**Results:** For histogram PCs, median ICCs for Cerebrum-7T, Freesurfer-v7, and Nighres were 0.99, 0.42, and 0.11 for the gray matter; 0.84, 0.25, and 0.43 for the basal ganglia; 0.89, 0.063, and 0.036 for the white matter; 0.84, 0.21, and 0.33 for the ventricles; 0.94, 0.64, and 0.93 for the cerebellum; and 0.78, 0.21, and 0.53 for the brainstem. For texture PCs, median ICCs for Cerebrum-7T, Freesurfer-v7, and Nighres were 0.95, 0.21, and 0.15 for the gray matter; 0.70, 0.36, and 0.023 for the basal ganglia; 0.91, 0.25, and 0.023 for the white matter; 0.80, 0.75, and 0.59 for the ventricles; 0.95, 0.43, and 0.86 for the cerebellum; and 0.72, 0.39, and 0.46 for the brainstem. For shape features, median ICCs for Cerebrum-7T, FreeSurfer-v7, and Nighres were 0.99, 0.91, and 0.36 for the gray matter; 0.89, 0.90, and 0.13 for the basal ganglia; 0.98, 0.91, and 0.027 for the white matter; 0.91, 0.91, and 0.36 for the ventricles; 0.80, 0.68, and 0.47 for the cerebellum; and 0.79, 0.17, and 0.15 for the brainstem.

**Conclusions:** Radiomic features in UHF MRI of the brain show substantial variability depending on the segmentation algorithm. The deep learning algorithm Cerebrum-7T enabled the highest reproducibility. Dedicated software tools for UHF MRI may be needed to achieve more stable results.

## INTRODUCTION

Ultrahigh-field (UHF) whole-body MRI scanners operating at 7T have recently been introduced into clinical practice following their initial CE certification. The higher field strength offers several advantages over standard 1.5 and 3T scanners, including increased spatial resolution and tissue contrast [1], which enables improved visualization of subcentimeter anatomic tissue properties. However, 7T MRI also has drawbacks, such as more pronounced artifacts at tissue-air interfaces (e.g., at the skull base and orbits) and secondary to motion, and substantial B0 and B1 field inhomogeneity [1].

Inhomogeneity and data size at 7T are also a problem for the application of segmentation tools that were developed using MR images obtained at lower field strengths. Such tools are now widely used in MRI research, for example to map functional MRI data to underlying brain structures. To address this problem in the area of neuroimaging, the deep-learning algorithm Cerebrum-7T has recently been proposed for fast and fully automatic segmentation of major brain structures on 7T images [2]. Cerebrum-7T showed good performance when compared to a partly automatic and partly manual, *de-facto* reference standard, and, in few cases, also to a purely manual segmentation. Cerebrum-7T also performed favorably compared to other commonly used segmentation tools [2].

The prior evaluation of Cerebrum-7T used three measures of segmentation similarity for evaluation of segmentation accuracy, with an emphasis on the Dice coefficient [2], but did not explore the impact of differences in segmentation masks on radiomic features at 7T that capture intra-volume properties such as signal heterogeneity. This topic is of interest because radiomic features –which are now widely used for the analysis of both focal brain lesions and diffuse abnormalities within the brain [3–9]– are sensitive to the type of segmentation method [10–14].

Based on the hypothesis that differences in segmentation masks would have a particularly strong effect on histogram-, texture-, and shape-based radiomic features at 7T, we reanalyzed the publicly accessible, held-out dataset that was used to test Cerebrum-7T. It was our aim to compare the reproducibility of radiomic features extracted from segmentation masks produced by Cerebrum-7T and two other segmentation tools, relative to partly manual reference standard segmentation.

## MATERIALS AND METHODS

### Subjects and design

We retrospectively analyzed 21 T1-weighted MRI scans of the brain from the Glasgow test dataset of the Cerebrum-7T study that are publicly available in the repository of the project (URL: https://rocknroll87q.github.io/cerebrum7t/). No Institutional Review Board or Ethics Committee approval was required as no patient identifiable information was accessed, and all data used for the analysis was sourced from a publicly available repository.

### MRI protocol

A 3D magnetization-prepared two rapid acquisition gradient echo (MP2RAGE) T1-weighted MRI sequence of the brain was obtained on a 7 Tesla MRI scanner (Siemens 7T Terra Magnetom) equipped with a 32-channel head coil, as previously described by Svanera et al [2]. Acquisition parameters were: repetition/echo time (TR/TE), 4680 ms/2.07 ms; inversion times (TI), 840 and 2370 ms; flip angles 5° and 6°; voxel size 0.63 x 0.625 x 0.625 mm³ with a base resolution was 384; FOV 204 x 220 x 178.5 mm. MP2RAGE is a commonly used sequence at 7T, because it is free of proton density and T2 contrasts and reduces magnetic field inhomogeneity effects while providing a high contrast-to-noise ratio [15]. The de-identified images are available for research purposes from https://rocknroll87q.github.io/cerebrum7t/data.

### Image analysis

We used the 3D Slicer software package (https://www.slicer.org/) version 5.4.0 [16] for all image analyses. Segmentations of six anatomic structures, as provided in the project repository –gray matter, basal ganglia, white matter, ventricles, cerebellum, and brainstem– which were obtained using the following four previously described segmentation techniques, were used:

1. *iGT (“inaccurate ground truth”):* A custom segmentation pipeline, used as the *de-facto* reference standard for the other three segmentation techniques. The iGT relied on two processing branches: one for white and gray matter segmentation, using a combination of the AFNI-3dSeg algorithm and a geometric and clustering technique [17, 18]; and one for specific segmentation of basal ganglia, ventricles, brainstem and cerebellum, using a combination of denoising and Freesurfer-v6 [19, 20]. Results of the two processing branches were then combined, and manual supervision and correction of the segmentations by an expert rater was performed as the final step, to ensure satisfactory results [2].
2. *Cerebrum-7T*: A fully automatic, deep-learning segmentation algorithm (deep encoder/decoder network with three layers) based on the original Cerebrum algorithm developed for 3T data [21]. Using the whole MRI volumes as network input, the convolutional neural network (CNN) was specifically trained on 7T MRI data (Glasgow training dataset), using the iGT segmentations as a reference standard [2]. The Cerebrum-7T code is available for download from https://github.com/rockNroll87q/cerebrum7t.
3. *Freesurfer-v7*: Version 7 of the Freesurfer image analysis suite, which is documented and freely available for download at http://surfer.nmr.mgh.harvard.edu and was utilized in a large number of neuroimaging studies, including several using 7T MRI [22–24]. Partly built on atlas information, this tool offers improvements for UHF image segmentation compared to prior versions [25].
4. *Nighres*: A Python-based toolbox building on the quantitative and high-resolution image-processing capabilities of the CBS High-Res Brain Processing Tools software suite, which was designed to specifically deal with submillimeter resolution MRI [26, 27]. Nighres is available for download from https://github.com/nighres/nighres.

Based on these segmentations, the PyRadiomics software module for 3D Slicer was used (available from https://github.com/AIM-Harvard/SlicerRadiomics) to extract a total of 101 three-dimensional radiomic features from each of the six brain regions, separately for each of the four segmentation algorithms: 18 based on the gray-level histogram; 74 texture features (including 24 from the co-occurrence matrix, 16 from the run-length matrix, 16 from the size-zone matrix, 13 from the gray-level dependence matrix, and 5 from the neighboring gray-tone difference matrix), and nine shape features (Elongation, Flatness, Maximum diameter, Major axis length, Minor axis length, Least axis length, Mesh volume, Sphericity, Surface area) [28]. The full list of features and equations is available at https://pyradiomics.readthedocs.io/en/latest/features.html. As preprocessing steps, intensity discretization using a fixed number of 64, and Z-score intensity normalization were performed. No spatial resampling was performed; the original isotropic voxel size of 0.63 mm³, which was identical for all included scans, was used.

### Statistical analysis

Given the large number of radiomic features, many of which may be highly correlated, dimensionality and data redundancy reduction by principal component analysis (PCA, based on Eigenvalues >1, with a maximum of 25 iterations for convergence) was applied, independently to the 18 gray-level histogram features and to the 74 texture features, across all brain structures and algorithms. No PCA was performed for the nine shape features. To test the reproducibility of radiomic features of the three classes, intraclass correlation coefficients (ICC) using a two-way mixed-effects model for absolute agreement were used to compare Cerebrum-7, Freesurfer-v7, and Nighres to the iGT, respectively. ICCs were interpreted as previously recommended: <0.5, poor; 0.5-0.75, moderate; 0.76-0.9, good; and >0.90, excellent reproducibility [29]. Analyses were performed using SPSS 28.0 (IBM Corp., Chicago, IL, USA).

## RESULTS

Feature extraction was successful for all segmentations (see Fig. 1). PCA identified five principal components (PC) for the gray-level histogram (originally 18 features), and seven PCs across all texture feature categories (originally 74 features).

**Figure 1.**
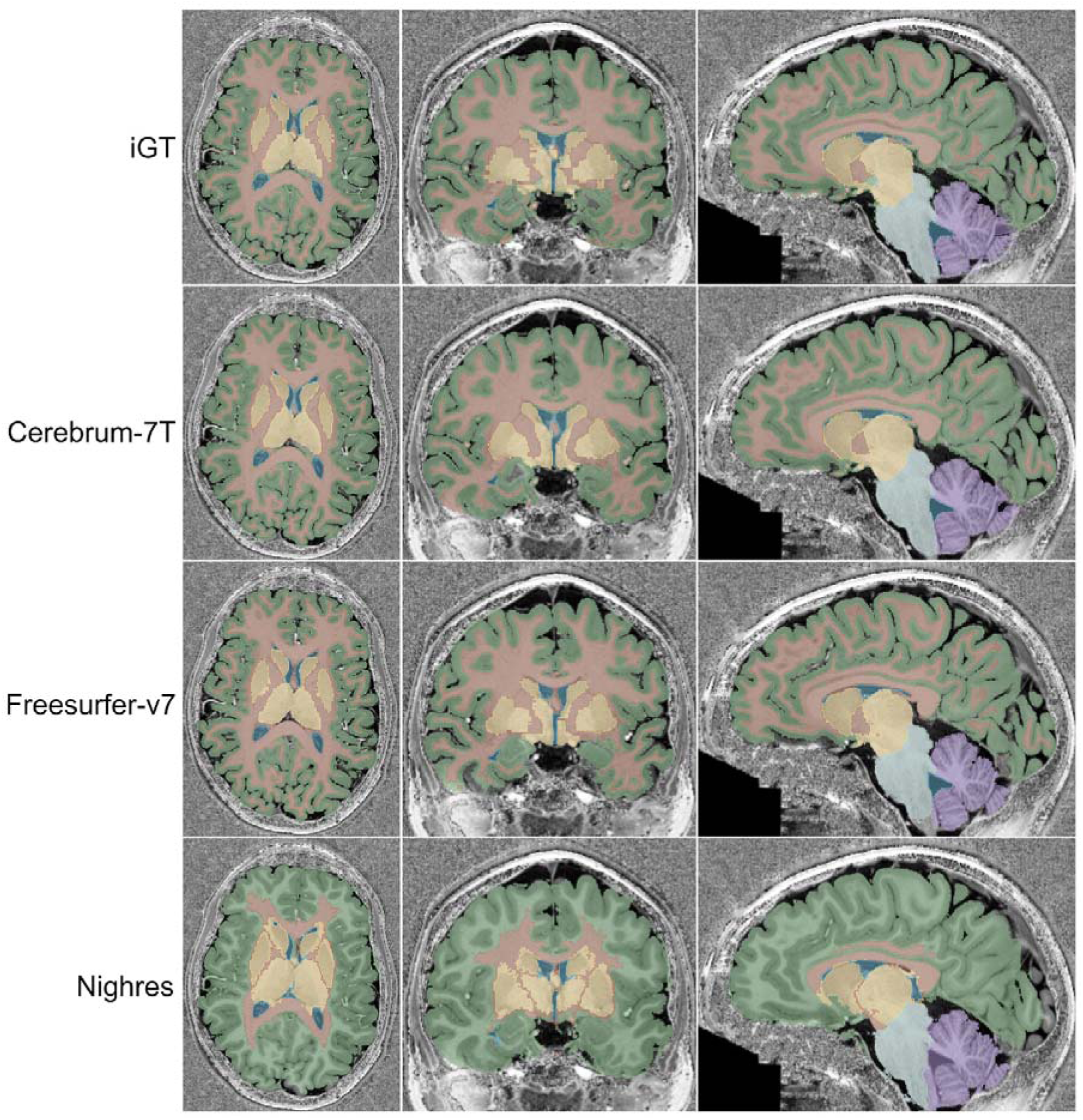
Comparison of the four segmentation techniques. While segmentation maps for gray matter (green), basal ganglia (yellow), white matter (brown), ventricles (blue), cerebellum (purple), and brainstem (light blue) were overall quite similar for the iGT, Cerebrum-7T and Freesurfer-v7 – although the inferior portion of the brainstem was missed by Freesurfer-v7– Nighres segmentation maps clearly differ, most prominently for the white matter.

Histogram and texture features extracted from the Cerebrum-7T segmentations generally showed clearly better reproducibility than those extracted from Freesurfer-v7 and Nighres segmentations, relative to features extracted from iGT segmentations (see Tables 1 and 2, Fig. 2). Reproducibility of features from Cerebrum-7T segmentations was excellent with ICCs >0.9 for 14/30 histogram PCs, and for 18/42 texture PCs (Tables 1 and 2); whereas it was poor (ICC<0.5) for only 2/42 texture PCs, and for none of the histogram PCs. Conversely, few histogram and texture PCs from Freesurfer-v7 and Nighres segmentations showed excellent, and many PCs, poor reproducibility, relative the iGT. For histogram PCs, median ICCs for Cerebrum-7T, Freesurfer-v7, and Nighres were 0.99, 0.42, and 0.11 for the gray matter; 0.84, 0.25, and 0.43 for the basal ganglia; 0.89, 0.063, and 0.036 for the white matter; 0.84, 0.21, and 0.33 for the ventricles; 0.94, 0.64, and 0.93 for the cerebellum; and 0.78, 0.21, and 0.53 for the brainstem. For texture PCs, median ICCs for Cerebrum-7T, Freesurfer-v7, and Nighres were 0.95, 0.21, and 0.15 for the gray matter; 0.70, 0.36, and 0.023 for the basal ganglia; 0.91, 0.25, and 0.023 for the white matter; 0.80, 0.75, and 0.59 for the ventricles; 0.95, 0.43, and 0.86 for the cerebellum; and 0.72, 0.39, and 0.46 for the brainstem.

**Figure 2.**
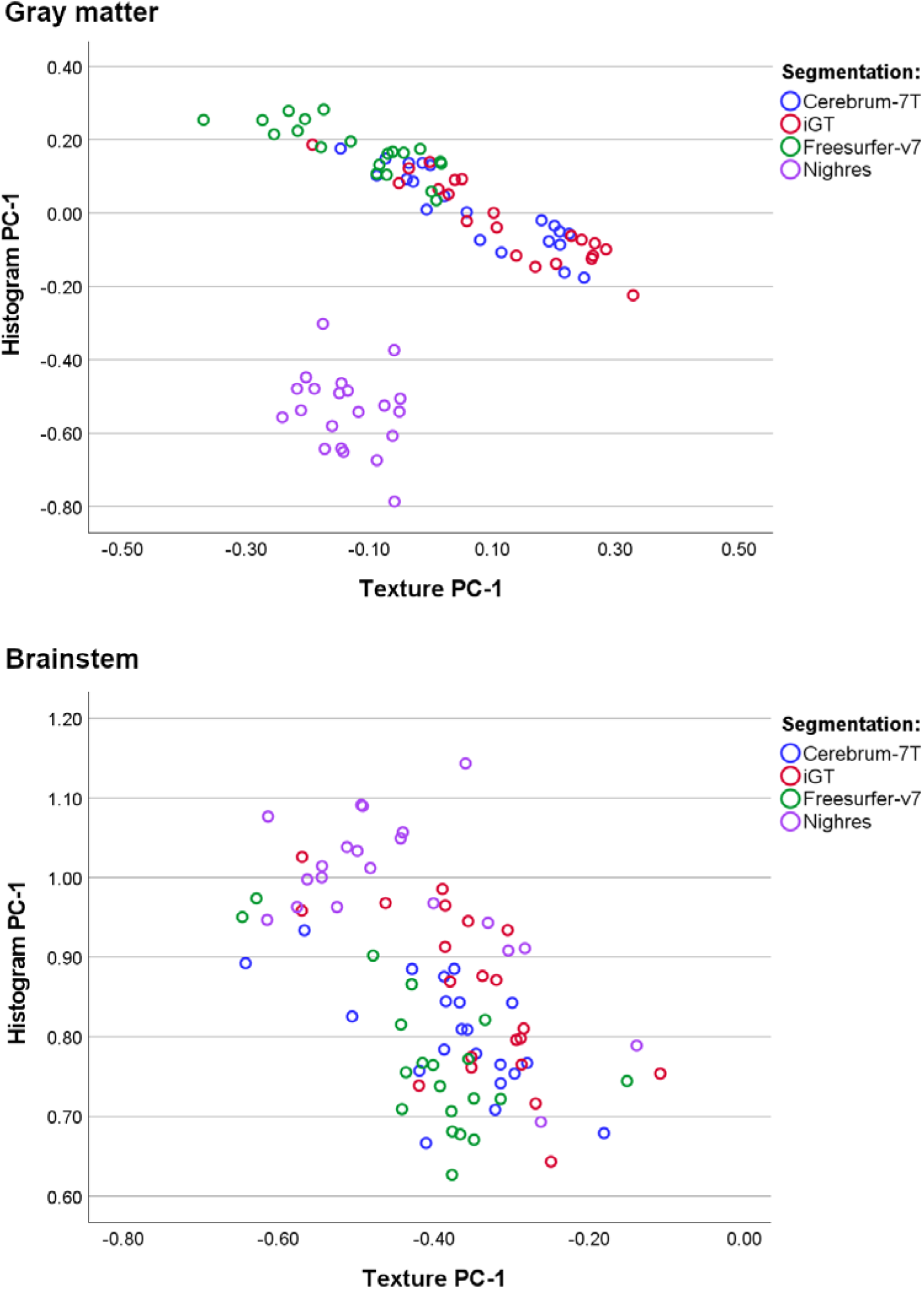
Exemplary scatter plots for two evaluated brain regions, based on the respective first histogram and texture principal components (PC). For the gray matter, Cerebrum-7T and iGT measurements show a considerable overlap, and a partial overlap with Freesurfer-v7 data, whereas the Nighres cluster is clearly separate. On the other hand, for the brainstem, there is less overlap between iGT measurements and those based on the other three segmentation techniques, including Cerebrum-7T.

**Table 1.** Histogram feature ICCs for the three segmentation algorithms relative to reference iGT.

|  | **Gray matter** | **Basal ganglia** | **White matter** | **Ventricles** | **Cerebellum** | **Brainstem** |
| --- | --- | --- | --- | --- | --- | --- |
| **Cerebrum-7T:** |  |  |  |  |  |  |
| PC-1 | 0.95 | 0.84* | 0.89* | 0.84* | 0.94* | 0.64 |
| PC-2 | 0.99 | 0.79 | 0.98 | 0.52 | 0.88 | 0.78* |
| PC-3 | 0.99* | 0.91 | 0.71 | 0.98 | 0.93 | 0.81 |
| PC-4 | 1.0 | 0.78 | 0.86 | 0.91 | 0.99 | 0.64 |
| PC-5 | 0.86 | 0.95 | 0.94 | 0.61 | 0.99 | 0.82 |
| **Freesurfer-v7:** |  |  |  |  |  |  |
| PC-1 | 0.15 | 0.19 | 0.022 | 0.14 | 0.47 | 0.46 |
| PC-2 | 0.22 | 0.66 | 0.081 | 0.43 | 0.79 | 0.56 |
| PC-3 | 0.91 | 0.25* | 0.34 | 0.81 | 0.89 | 0.062 |
| PC-4 | 0.42* | 0.42 | 0.28 | 0.21* | 0.40 | 0.21* |
| PC-5 | -0.022 | 0.036 | 0.063* | 0.14 | 0.64* | 0.17 |
| **Nighres:** |  |  |  |  |  |  |
| PC-1 | 0.066 | 0.49 | -0.065 | 0.33* | 0.88 | 0.005 |
| PC-2 | 0.19 | 0.082 | 0.036* | 0.051 | 0.84 | 0.84 |
| PC-3 | 0.11* | 0.43* | -0.002 | 0.53 | 0.93* | -0.23 |
| PC-4 | 0.75 | 0.89 | 0.11 | 0.66 | 0.98 | 0.68 |
| PC-5 | 0.021 | 0.19 | 0.047 | 0.14 | 0.95 | 0.53* |

\* Median within each anatomic region

**Table 2.** Texture feature ICCs for the three segmentation algorithms relative to reference iGT.

|  | **Gray**<br>**matter** | **Basal**<br>**ganglia** | **White**<br>**matter** | **Ventricles** | **Cerebellum** | **Brainstem** |
| --- | --- | --- | --- | --- | --- | --- |
| **Cerebrum-7T:** |  |  |  |  |  |  |
| PC-1 | 0.91 | 0.93 | 0.70 | 0.63 | 0.95* | 0.87 |
| PC-2 | 0.99 | 0.70* | 0.82 | 0.93 | 0.73 | 0.79 |
| PC-3 | 0.95* | 0.72 | 0.99 | 0.80* | 0.80 | 0.72* |
| PC-4 | 0.38 | 0.91 | 0.97 | 0.48 | 0.99 | 0.57 |
| PC-5 | 0.98 | 0.66 | 0.91* | 0.64 | 0.74 | 0.70 |
| PC-6 | 0.92 | 0.67 | 0.68 | 0.86 | 0.96 | 0.71 |
| PC-7 | 0.97 | 0.53 | 0.96 | 0.91 | 0.97 | 0.85 |
| **Freesurfer-v7:** |  |  |  |  |  |  |
| PC-1 | 0.21* | 0.35 | 0.13 | 0.48 | 0.43* | 0.39* |
| PC-2 | 0.19 | 0.19 | -0.048 | 0.94 | 0.021 | 0.30 |
| PC-3 | 0.54 | 0.73 | 0.62 | 0.53 | 0.65 | 0.49 |
| PC-4 | -0.027 | 0.72 | 0.25* | 0.20 | 0.57 | 0.14 |
| PC-5 | 0.52 | 0.52 | 0.96 | 0.75* | 0.27 | 0.40 |
| PC-6 | 0.12 | 0.36* | -0.048 | 0.75 | 0.67 | 0.48 |
| PC-7 | 0.61 | 0.26 | 0.36 | 0.94 | 0.18 | 0.32 |
| **Nighres:** |  |  |  |  |  |  |
| PC-1 | 0.099 | 0.27 | 0.12 | 0.59* | 0.91 | 0.29 |
| PC-2 | 0.51 | 0.20 | 0.023* | 0.80 | 0.55 | 0.46* |
| PC-3 | 0.71 | -0.025 | -0.008 | 0.50 | 0.86* | 0.51 |
| PC-4 | 0.0001 | 0.67 | -0.064 | 0.013 | 0.98 | 0.31 |
| PC-5 | 0.069 | 0.001 | 0.098 | 0.46 | 0.73 | 0.43 |
| PC-6 | 0.15* | -0.012 | -0.11 | 0.89 | 0.95 | 0.52 |
| PC-7 | 0.74 | 0.023* | 0.024 | 0.81 | 0.70 | 0.51 |

\* Median within each anatomic region

Shape features extracted from the Cerebrum-7T segmentations again showed the overall best reproducibility relative to the iGT, but differences to the two other segmentation techniques were less pronounced, especially to Freesurfer-v7, which showed better reproducibility for several individual features (Table 3). For the feature Sphericity, reproducibility was poor for all three segmentation tools. Median shape feature ICCs for Cerebrum-7T, FreeSurfer-v7, and Nighres were 0.99, 0.91, and 0.36 for the gray matter; 0.89, 0.90, and 0.13 for the basal ganglia; 0.98, 0.91, and 0.027 for the white matter;; 0.91, 0.91, and 0.36 for the ventricles; 0.80, 0.68, and 0.47 for the cerebellum; and 0.79, 0.17, and 0.15 for the brainstem.

**Table 3.**
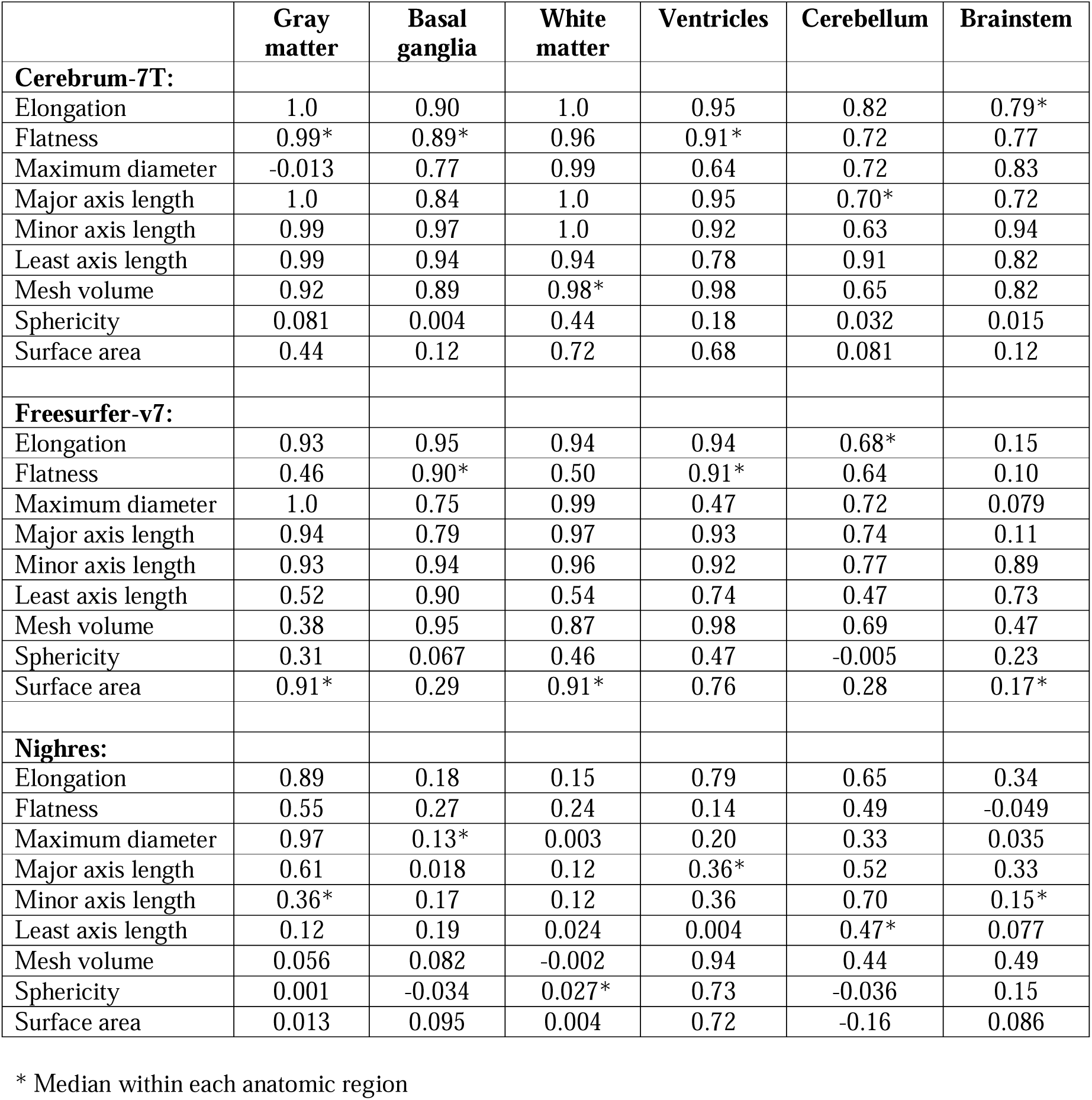
Shape feature ICCs for the three segmentation algorithms relative to reference iGT.

|  | **Gray matter** | **Basal ganglia** | **White matter** | **Ventricles** | **Cerebellum** | **Brainstem** |
| --- | --- | --- | --- | --- | --- | --- |
| **Cerebrum-7T:** |  |  |  |  |  |  |
| Elongation | 1.0 | 0.90 | 1.0 | 0.95 | 0.82 | 0.79* |
| Flatness | 0.99* | 0.89* | 0.96 | 0.91* | 0.72 | 0.77 |
| Maximum diameter | -0.013 | 0.77 | 0.99 | 0.64 | 0.72 | 0.83 |
| Major axis length | 1.0 | 0.84 | 1.0 | 0.95 | 0.70* | 0.72 |
| Minor axis length | 0.99 | 0.97 | 1.0 | 0.92 | 0.63 | 0.94 |
| Least axis length | 0.99 | 0.94 | 0.94 | 0.78 | 0.91 | 0.82 |
| Mesh volume | 0.92 | 0.89 | 0.98* | 0.98 | 0.65 | 0.82 |
| Sphericity | 0.081 | 0.004 | 0.44 | 0.18 | 0.032 | 0.015 |
| Surface area | 0.44 | 0.12 | 0.72 | 0.68 | 0.081 | 0.12 |
| **Freesurfer-v7:** |  |  |  |  |  |  |
| Elongation | 0.93 | 0.95 | 0.94 | 0.94 | 0.68* | 0.15 |
| Flatness | 0.46 | 0.90* | 0.50 | 0.91* | 0.64 | 0.10 |
| Maximum diameter | 1.0 | 0.75 | 0.99 | 0.47 | 0.72 | 0.079 |
| Major axis length | 0.94 | 0.79 | 0.97 | 0.93 | 0.74 | 0.11 |
| Minor axis length | 0.93 | 0.94 | 0.96 | 0.92 | 0.77 | 0.89 |
| Least axis length | 0.52 | 0.90 | 0.54 | 0.74 | 0.47 | 0.73 |
| Mesh volume | 0.38 | 0.95 | 0.87 | 0.98 | 0.69 | 0.47 |
| Sphericity | 0.31 | 0.067 | 0.46 | 0.47 | -0.005 | 0.23 |
| Surface area | 0.91* | 0.29 | 0.91* | 0.76 | 0.28 | 0.17* |
| **Nighres:** |  |  |  |  |  |  |
| Elongation | 0.89 | 0.18 | 0.15 | 0.79 | 0.65 | 0.34 |
| Flatness | 0.55 | 0.27 | 0.24 | 0.14 | 0.49 | -0.049 |
| Maximum diameter | 0.97 | 0.13* | 0.003 | 0.20 | 0.33 | 0.035 |
| Major axis length | 0.61 | 0.018 | 0.12 | 0.36* | 0.52 | 0.33 |
| Minor axis length | 0.36* | 0.17 | 0.12 | 0.36 | 0.70 | 0.15* |
| Least axis length | 0.12 | 0.19 | 0.024 | 0.004 | 0.47* | 0.077 |
| Mesh volume | 0.056 | 0.082 | -0.002 | 0.94 | 0.44 | 0.49 |
| Sphericity | 0.001 | -0.034 | 0.027* | 0.73 | -0.036 | 0.15 |
| Surface area | 0.013 | 0.095 | 0.004 | 0.72 | -0.16 | 0.086 |

\* Median within each anatomic region

## DISCUSSION

The results of our study demonstrate that the fully automatic Cerebrum-7T deep-learning segmentation algorithm, specifically developed for 7T data, achieved the overall highest reproducibility of radiomic features across the different brain regions, relative to the iGT. This was especially true for histogram and texture features, and most prominently visible in the cerebral gray and white matter as well as the cerebellum, where many PCs showed ICCs of >0.9 (see Tables 1 and 2). Contrary to that, both Freesurfer-v7 and Nighres performed poorly for the majority of evaluated PCs, and across all segmented brain regions. While this is not particularly surprising for Nighres, for which Svanera et al. reported low Dice coefficients especially for the gray and white matter, it is quite surprising for Freesurfer-v7, which had previously demonstrated good segmentation performance for most brain regions [2]. These results provide further proof that, within the radiomics workflow, segmentation is a critical step where minor differences in segmentation masks can have a profound effect on calculated feature values. This is also underscored by the fact that even Cerebrum-7T, although trained directly on iGT segmentations, showed only moderate-to-good histogram and texture feature reproducibility in the brainstem, and partly also in the ventricles (although the clinical value of signal characteristics obtained from the ventricles is probably limited).

Shape feature results, on the other hand, differed in several aspects from the above-described histogram and texture feature results. While Nighres again performed quite poorly for the majority of features, with only few exceptions, feature reproducibility differences between Cerebrum-7T and Freesurfer-v7 segmentations were less pronounced, and for some features and anatomic regions, minimal or nonexistent (see Table 3). Axis length measurements, which may also be used clinically, for example to assess cortical thickness, showed mostly good to excellent reproducibility across brain regions for both Cerebrum-7T and Freesurfer-v7. Only for the brainstem was Cerebrum-7T clearly superior to both Freesurfer-v7 (and also Nighres). Surprisingly, for the feature Sphericity, which is a descriptor of roundness of the segmented shape relative to a sphere, there was practically no correlation between the iGT and all three segmentation methods evaluated, the reason for which is unclear.

Svanera et al. originally used three metrics, which were previously recommended in the MICCAI MRBrainS18 challenge to benchmark segmentation algorithms, for comparison of Cerebrum-7T to the reference standard: the Dice coefficient to assess the overlap between segmentations; the 95th percentile of the Hausdorff Distance to assess the proximity between segmentation contours; and the Volumetric Similarity as a non-overlap-based metric to assess volumetric similarity [30]. While these well-established metrics capture the quality of the segmentation, they do not assess quantitative differences between segmentations in terms of voxel-based gray-level statistics and patterns contained therein. This information, which is captured by radiomic features, is currently evaluated in a wide array of diseases of the brain, with recent MRI applications including detection of cognitive impairment [8], distinguishing between active and chronic multiple myeloma lesions [9], EGFR and HER2 status prediction in adenocarcinoma brain metastases [5], corticospinal tract involvement in glioma [4], and prediction of local tumor control in patients with brain metastases following postoperative radiotherapy [6].

Previous studies have demonstrated that the results of radiomics-based classification are generally improved at higher spatial resolution [31, 32], which is one of the key strengths of UHF MRI over MRI at lower field strengths. However, voxel intensity inhomogeneity and artifacts at 7T may negatively impact feature values, and in particular, their reproducibility. Based on prior MRI research in different areas of the body, including the brain, which already showed that radiomic feature are sensitive to variations in segmentation [10–14], we hypothesized that differences in segmentation masks would have a particularly strong impact of radiomic features at 7T. Such major differences were indeed observed in our study, in particular for segmentation tools that were not optimized for the specific type of MRI dataset.

Our study has several limitations, including first the use of the iGT as a reference standard. The reason is that Cerebrum-7T was trained using the iGT –although on a different dataset that was not used in this study– and that therefore, Cerebrum-7T segmentations may more closely resemble those of the iGT than segmentations created by the two other methods, which were independent of the iGT. On the other hand, the iGT included a manual, human expert-level correction step at the end, and therefore, was the logical choice of reference standard out of the available segmentation techniques. Another limitation was our inability to directly compare the reproducibility data at 7T to a matching dataset obtained at 3T or 1.5T, because no such data was available in the repository. Finally, the analysis only included a single, T1-weighted MRI pulse sequence, whereas clinical as well as research protocols also include other sequences, such as fluid-attenuated inversion recovery (FLAIR) and/or T2-weighted sequences.

In conclusion, the results of our study demonstrate that the deep learning-based segmentation algorithm Cerebrum-7T, which was specifically trained on 7T data, provided a higher degree of radiomic feature reproducibility than other available segmentation tools. These findings support the notion that, for the processing of UHF MRI data, dedicated analysis tools such as segmentation algorithms may need to be developed, especially since 7T MR scanners have now entered the clinical stage. Our results also provide further evidence that histogram, texture, as well as shape features are highly sensitive to the choice of segmentation technique, and comparison between results of different studies must take this factor into account.

## Data Availability

All data produced in the present study are available upon reasonable request to the authors

https://search.kg.ebrains.eu/instances/Dataset/2b24466d-f1cd-4b66-afa8-d70a6755ebea

## Notes

Conflicts of Interest and Source of Funding: M.E.M. received honoraria for lectures from Siemens, General Electric, and Bristol Myers Squibb. The other authors declare no potential conflicts of interest.

### Competing Interest Statement

M.E.M. received honoraria for lectures from Siemens, General Electric, and Bristol Myers Squibb. The other authors declare no potential conflicts of interest.

### Funding Statement

This study did not receive any funding

### Author Declarations

The study used (or will use) ONLY openly available human data that were originally located at: https://search.kg.ebrains.eu/instances/Dataset/2b24466d-f1cd-4b66-afa8-d70a6755ebea

## REFERENCES

1. Okada T, Fujimoto K, Fushimi Y, Akasaka T, Thuy DHD, Shima A, Sawamoto N, Oishi N, Zhang Z, Funaki T, Nakamoto Y, Murai T, Miyamoto S, Takahashi R, Isa T. Neuroimaging at 7 Tesla: a pictorial narrative review. Quant Imaging Med Surg. 2022 Jun;12(6):3406–3435. doi: 10.21037/qims-21-969.

2. Svanera M, Benini S, Bontempi D, Muckli L. CEREBRUM-7T: Fast and Fully Volumetric Brain Segmentation of 7 Tesla MR Volumes. Hum Brain Mapp. 2021 Dec 1;42(17):5563–5580. doi: 10.1002/hbm.25636.

3. Nenning KH, Gesperger J, Furtner J, Nemc A, Roetzer-Pejrimovsky T, Choi SW, Mitter C, Leber SL, Hofmanninger J, Klughammer J, Ergüner B, Bauer M, Brada M, Chong K, Brandner-Kokalj T, Freyschlag CF, Grams A, Haybaeck J, Hoenigschnabl S, Hoffermann M, Iglseder S, Kiesel B, Kitzwoegerer M, Kleindienst W, Marhold F, Moser P, Oberndorfer S, Pinggera D, Scheichel F, Sherif C, Stockhammer G, Stultschnig M, Thomé C, Trenkler J, Urbanic-Purkart T, Weis S, Widhalm G, Wuertz F, Preusser M, Baumann B, Simonitsch-Klupp I, Nam DH, Bock C, Langs G, Woehrer A. Radiomic features define risk and are linked to DNA methylation attributes in primary CNS lymphoma. Neurooncol Adv. 2023 Oct 18;5(1):vdad136. doi: 10.1093/noajnl/vdad136.

4. Wende T, Güresir E, Wach J, Vychopen M, Hoffmann A, Prasse G, Wilhelmy F, Kasper J. Radiomic white matter parameters of functional integrity of the corticospinal tract in high-grade glioma. Sci Rep. 2024 Jun 5;14(1):12891. doi: 10.1038/s41598-024-63813-2.

5. Li Y, Jin Y, Wang Y, Liu W, Jia W, Wang J. MR-based radiomics predictive modelling of EGFR mutation and HER2 overexpression in metastatic brain adenocarcinoma: a two-centre study. Cancer Imaging. 2024 May 21;24(1):65. doi: 10.1186/s40644-024-00709-4.

6. Buchner JA, Kofler F, Mayinger M, Christ SM, Brunner TB, Wittig A, Menze B, Zimmer C, Meyer B, Guckenberger M, Andratschke N, El Shafie RA, Debus J, Rogers S, Riesterer O, Schulze K, Feldmann HJ, Blanck O, Zamboglou C, Ferentinos K, Bilger-Zähringer A, Grosu AL, Wolff R, Piraud M, Eitz KA, Combs SE, Bernhardt D, Rueckert D, Wiestler B, Peeken JC. Radiomics-based prediction of local control in patients with brain metastases following postoperative stereotactic radiotherapy. Neuro Oncol. 2024 May 30:noae098. doi: 10.1093/neuonc/noae098.

7. Shu Z, Pang P, Wu X, Cui S, Xu Y, Zhang M. An Integrative Nomogram for Identifying Early-Stage Parkinson’s Disease Using Non-motor Symptoms and White Matter-Based Radiomics Biomarkers From Whole-Brain MRI. Front Aging Neurosci. 2020 Dec 17;12:548616. doi: 10.3389/fnagi.2020.548616.

8. Feng J, Hui D, Zheng Q, Guo Y, Xia Y, Shi F, Zhou Q, Yu F, He X, Wang S, Li C. Automatic detection of cognitive impairment in patients with white matter hyperintensity and causal analysis of related factors using artificial intelligence of MRI. Comput Biol Med. 2024 Jun 4;178:108684. doi: 10.1016/j.compbiomed.2024.108684.

9. Caba B, Cafaro A, Lombard A, Arnold DL, Elliott C, Liu D, Jiang X, Gafson A, Fisher E, Belachew SM, Paragios N. Single-timepoint low-dimensional characterization and classification of acute versus chronic multiple sclerosis lesions using machine learning. Neuroimage. 2023 Jan;265:119787. doi: 10.1016/j.neuroimage.2022.119787.

10. Saha A, Harowicz MR, Mazurowski MA. Breast cancer MRI radiomics: An overview of algorithmic features and impact of inter-reader variability in annotating tumors. Med Phys. 2018 Jul;45(7):3076–3085.

11. Tixier F, Um H, Young RJ, Veeraraghavan H. Reliability of tumor segmentation in glioblastoma: Impact on the robustness of MRI-radiomic features. Med Phys. 2019 Aug;46(8):3582–3591. doi: 10.1002/mp.13624.

12. Granzier RWY, Verbakel NMH, Ibrahim A, van Timmeren JE, van Nijnatten TJA, Leijenaar RTH, Lobbes MBI, Smidt ML, Woodruff HC. MRI-based radiomics in breast cancer: feature robustness with respect to inter-observer segmentation variability. Sci Rep. 2020 Aug 25;10(1):14163. doi: 10.1038/s41598-020-70940-z.

13. Chen H, Li S, Zhang Y, Liu L, Lv X, Yi Y, Ruan G, Ke C, Feng Y. Deep learning-based automatic segmentation of meningioma from multiparametric MRI for preoperative meningioma differentiation using radiomic features: a multicentre study. Eur Radiol. 2022 Oct;32(10):7248–7259. doi: 10.1007/s00330-022-08749-9.

14. Poirot MG, Caan MWA, Ruhe HG, Bjørnerud A, Groote I, Reneman L, Marquering HA. Robustness of radiomics to variations in segmentation methods in multimodal brain MRI. Sci Rep. 2022 Oct 6;12(1):16712. doi: 10.1038/s41598-022-20703-9.

15. Marques JP, Kober T, Krueger G, van der Zwaag W, Van de Moortele PF, Gruetter R. MP2RAGE, a self bias-field corrected sequence for improved segmentation and T1-mapping at high field. Neuroimage. 2010 Jan 15;49(2):1271–81. doi: 10.1016/j.neuroimage.2009.10.002.

16. Fedorov A, Beichel R, Kalpathy-Cramer J, Finet J, Fillion-Robin JC, Pujol S, Bauer C, Jennings D, Fennessy F, Sonka M, Buatti J, Aylward S, Miller JV, Pieper S, Kikinis R. 3D Slicer as an image computing platform for the Quantitative Imaging Network. Magn Reson Imaging. 2012 Nov;30(9):1323–41. doi: 10.1016/j.mri.2012.05.001.

17. Cox RW. AFNI: software for analysis and visualization of functional magnetic resonance neuroimages. Comput Biomed Res. 1996 Jun;29(3):162–73.

18. Desikan RS, Ségonne F, Fischl B, Quinn BT, Dickerson BC, Blacker D, Buckner RL, Dale AM, Maguire RP, Hyman BT, Albert MS, Killiany RJ. An automated labeling system for subdividing the human cerebral cortex on MRI scans into gyral based regions of interest. Neuroimage. 2006 Jul 1;31(3):968–80.

19. Fischl B. FreeSurfer. Neuroimage. 2012 Aug 15;62(2):774–81.

20. O’Brien KR, Kober T, Hagmann P, Maeder P, Marques J, Lazeyras F, Krueger G, Roche A. Robust T1-weighted structural brain imaging and morphometry at 7T using MP2RAGE. PLoS One. 2014 Jun 16;9(6):e99676.

21. Bontempi D, Benini S, Signoroni A, Svanera M, Muckli L. CEREBRUM: a fast and fully-volumetric Convolutional Encoder-decodeR for weakly-supervised sEgmentation of BRain strUctures from out-of-the-scanner MRI. Med Image Anal. 2020 May;62:101688.

22. Mueller SG. 7T MP2RAGE for cortical myelin segmentation: Impact of aging. PLoS One. 2024 Apr 16;19(4):e0299670. doi: 10.1371/journal.pone.0299670.

23. Lyu J, Bartlett PF, Nasrallah FA, Tang X. Toward hippocampal volume measures on ultra-high field magnetic resonance imaging: a comprehensive comparison study between deep learning and conventional approaches. Front Neurosci. 2023 Dec 14;17:1238646. doi: 10.3389/fnins.2023.1238646.

24. Choi EY, Tian L, Su JH, Radovan MT, Tourdias T, Tran TT, Trelle AN, Mormino E, Wagner AD, Rutt BK. Thalamic nuclei atrophy at high and heterogenous rates during cognitively unimpaired human aging. Neuroimage. 2022 Nov 15;262:119584. doi: 10.1016/j.neuroimage.2022.119584.

25. Zaretskaya N, Fischl B, Reuter M, Renvall V, Polimeni JR. Advantages of cortical surface reconstruction using submillimeter 7 T MEMPRAGE. Neuroimage. 2018 Jan 15;165:11–26.

26. Huntenburg JM, Steele CJ, Bazin PL. Nighres: processing tools for high-resolution neuroimaging. Gigascience. 2018 Jul 1;7(7):giy082.

27. Bazin PL, Weiss M, Dinse J, Schäfer A, Trampel R, Turner R. A computational framework for ultra-high resolution cortical segmentation at 7Tesla. Neuroimage. 2014 Jun;93 Pt 2:201–9.

28. van Griethuysen JJM, Fedorov A, Parmar C, Hosny A, Aucoin N, Narayan V, Beets-Tan RGH, Fillion-Robin JC, Pieper S, Aerts HJWL. Computational Radiomics System to Decode the Radiographic Phenotype. Cancer Res. 2017 Nov 1;77(21):e104–e107. doi: 10.1158/0008-5472.CAN-17-0339.

29. Koo TK, Li MY. A Guideline of Selecting and Reporting Intraclass Correlation Coefficients for Reliability Research. J Chiropr Med. 2016 Jun;15(2):155–63. doi: 10.1016/j.jcm.2016.02.012. Epub 2016 Mar 31. Erratum in: J Chiropr Med. 2017 Dec;16(4):346. doi: 10.1016/j.jcm.2017.10.001.

30. Taha AA, Hanbury A. Metrics for evaluating 3D medical image segmentation: analysis, selection, and tool. BMC Med Imaging. 2015 Aug 12;15:29. doi: 10.1186/s12880-015-0068-x.

31. Mayerhoefer ME, Szomolanyi P, Jirak D, Materka A, Trattnig S. Effects of MRI acquisition parameter variations and protocol heterogeneity on the results of texture analysis and pattern discrimination: an application-oriented study. Med Phys. 2009 Apr;36(4):1236–43. doi: 10.1118/1.3081408.

32. Wichtmann BD, Harder FN, Weiss K, Schönberg SO, Attenberger UI, Alkadhi H, Pinto Dos Santos D, Baeßler B. Influence of Image Processing on Radiomic Features From Magnetic Resonance Imaging. Invest Radiol. 2023 Mar 1;58(3):199–208. doi: 10.1097/RLI.0000000000000921.

